## Supplementary Text for "The effect of COVID-19 vaccination in Italy and perspectives for “living with the virus”"

### 1. Materials and Methods

#### 1.1 Model for SARS-CoV-2 transmission and vaccination

We developed an age-structured stochastic model for SARS-CoV-2 transmission and vaccination, based on a susceptible-infectious-removed-susceptible scheme (SIRS) and adapted from previously published models [1,2]. Mixing patterns are assumed to be heterogeneous across ages according to an age-specific social contact matrix estimated prior to the COVID-19 pandemic [3]. We assume an age-dependent susceptibility to SARS-CoV-2 infection: lower in children under 15 years of age and higher for the elderly (65+), compared to individuals of working age [4].

We simulate a two-dose vaccination campaign. In the baseline analysis, vaccination is assumed to reduce the individuals' susceptibility to SARS-CoV-2 infection and the risk of death. Breakthrough infections (i.e., infections in vaccinated individuals) are assumed to be half as infectious as those in unvaccinated individuals [5,6]. The model accounts for waning of both natural and vaccine-induced protection, where the duration is exponentially distributed (mean: 2 years [7,8]). Specifically, before waning of natural immunity individuals are fully protected against infection; while before waning of vaccine protection, vaccine reduces the probability of infection and death (with different efficacy estimates for the two endpoints) and the infectiousness of breakthrough infections. After waning of either natural or vaccine protection, individuals are considered fully susceptible. Alternative durations for natural immunity and vaccine protection are explored as sensitivity analyses (1 year and 10 years), as well as the case of a vaccine not reducing infectiousness (see Section 2.1). Immunity gained after experiencing both vaccination and infection (independently of the order) is assumed not to wane over the time scale of our simulations.

The baseline model is described by the following system of differential equations (summarized by the schematic representation in Figure S1):

$$\begin{cases} S'_{a,c}(t) = -\lambda_a(t)S_{a,c}(t) - \alpha_{a,c}(t)S_{a,c}(t) + \upsilon_N R_{a,c}(t) \\ I'_{a,c}(t) = \lambda_a(t)S_{a,c}(t) - \gamma I_{a,c}(t) - \alpha_{a,c}(t)I_{a,c}(t) \\ R'_{a,c}(t) = \gamma I_{a,c}(t) - \alpha_{a,c}(t)R_{a,c}(t) - \upsilon_N R_{a,c}(t) \\ U'_{a,c}(t) = \alpha_{a,c}(t)(I_{a,c}(t) + R_{a,c}(t)) \\ V'_{1,a,c}(t) = \alpha_{a,c}(t)S_{a,c}(t) - (1 - VE_{1,a}^{inf})\lambda_a(t)V_{1,a,c} - \omega_0 V_{1,a,c}(t) \\ V'_{2,a,c}(t) = \omega_0 V_{1,a,c}(t) - (1 - VE_{2,a}^{inf})\lambda_a(t)V_{2,a,c} - \omega_1 V_{2,a,c}(t) \\ V'_{3,a,c}(t) = \omega_1 V_{2,a,c}(t) - (1 - VE_{3,a}^{inf})\lambda_a(t)V_{3,a,c} - \omega_2 V_{3,a,c}(t) \\ V'_{4,a,c}(t) = \omega_2 V_{3,a,c}(t) - (1 - VE_{4,a}^{inf})\lambda_a(t)V_{4,a,c} - \upsilon_V V_{4,a,c}(t) \\ V'_{5,a,c}(t) = \upsilon_V V_{4,a,c}(t) - (1 - VE_{5,a}^{inf})\lambda_a(t)V_{5,a,c}(t) \\ I^{V\,\prime}_{a,c}(t) = \lambda_a(t)[(1 - VE_{1,a}^{inf})V_{1,a,c} + (1 - VE_{2,a}^{inf})V_{2,a,c} + (1 - VE_{3,a}^{inf})V_{3,a,c} + (1 - VE_{4,a}^{inf})V_{4,a,c} + (1 - VE_{5,a}^{inf})V_{5,a,c}(t)] - \gamma I^V_{a,c}(t) \\ R^{V\,\prime}_{a,c}(t) = \gamma I^V_{a,c}(t) \end{cases}$$

where:

- the population class {a,c} represents individuals in age group a and in underlying conditions status c ("with" or "without");
- $S_{a,c}$ represents the number of unvaccinated individuals in the population class {a,c} who are fully susceptible to SARS-CoV-2 infection;
- $I_{a,c}$ represents the number of infectious unvaccinated individuals in the population class {a,c}.
- $R_{a,c}$ represents the number of unvaccinated individuals in the population class {a,c} who recovered from infection.
- $U_{a,c}$ represents the number of individuals in the population class {a,c} who are vaccinated despite having already experienced SARS-CoV-2 infection.
- $V_{1,a,c}$; $V_{2,a,c}$; $V_{3,a,c}$; $V_{4,a,c}$ and $V_{5,a,c}$ represent the number of vaccinated individuals at different stages of protection. In particular,
    1) $V_{1,a,c}$ denotes individuals in the population class {a,c} vaccinated with the first dose, for whom the first dose is not effective yet.
    2) $V_{2,a,c}$ denotes individuals in the population class {a,c} vaccinated with the first dose, for whom the first dose is effective.
    3) $V_{3,a,c}$ denotes individuals in the population class {a,c} vaccinated with the second dose for whom the second dose is not effective yet.

4) $V_{4,a,c}$ denotes individuals in the population class {a,c} vaccinated with the second dose for whom the second dose is effective.
5) $V_{5,a,c}$ denotes individuals in the population class {a,c} vaccinated with the second dose for whom vaccine protection has waned.

- $I^V_{a,c}$ represents the number of infectious individuals in the population class {a,c} among those who have already received at least one vaccine dose.
- $R^V_{a,c}$ represents the number of individuals in the population class {a,c} who recovered from an infection contracted after having received at least one vaccine dose.

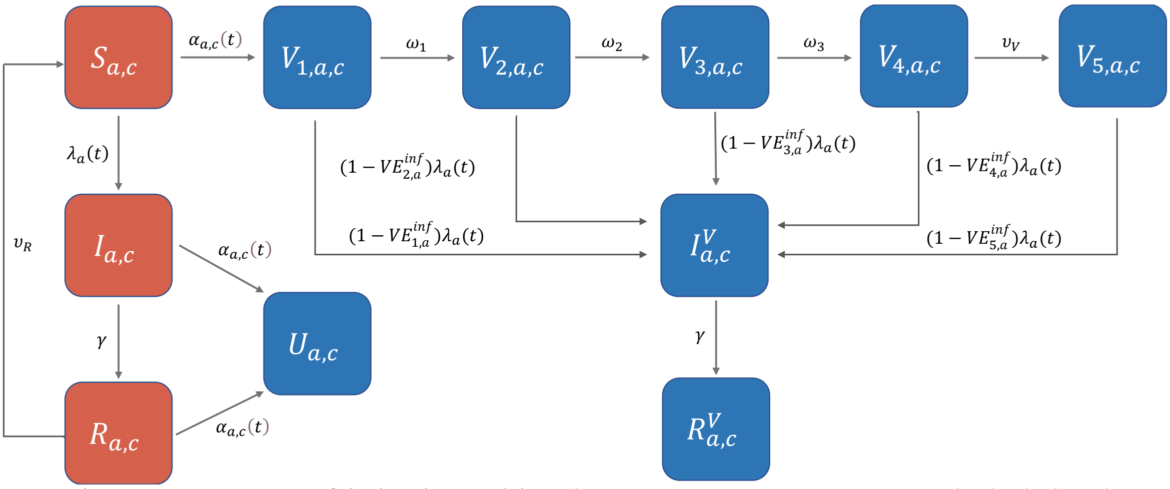

**Figure S1. Schematic representation of the baseline model.** Red compartments represent unvaccinated individuals and therefore eligible for vaccination; blue compartments represent vaccinated individuals. Model parameters include: the time- and age-dependent force of infection $\lambda_a(t)$; the recovery rate from infection ($\gamma$); the duration of immunity after infection ($1/\upsilon_R$); the time-, age- and group-dependent vaccination rate $\alpha_{a,c}(t)$; the average interval between administration of the first dose and full protection by the first dose ($1/\omega_1$); the average interval between full protection of the first dose and administration of the second dose ($1/\omega_2$); the average interval between administration of the second dose and full protection of the 2nd dose ($1/\omega_3$); the average duration of vaccine protection after administration of the second dose ($1/\upsilon_V$); age-dependent vaccine efficacies against infection in the different stages *i* of vaccine protection (i=1,2,3,4,5) are denoted by $VE^{inf}_{i,a}$.

Susceptible individuals are exposed to a time and age-dependent force of infection $\lambda_a(t)$ which is defined as:

$$\lambda_a(t) = \beta(1+\theta_{\text{Alpha}})(1-\varphi)\,\delta(t) r_a \sum_{\tilde{a}} C_{a,\tilde{a}} \frac{\sum_c \left[I_{\tilde{a},c}(t) + \pi I^V_{\tilde{a},c}(t)\right]}{\sum_c N_{\tilde{a},c}}$$

where:

- $\beta$ is a scaling factor shaping SARS-CoV-2 transmissibility in the absence of physical distancing restrictions (PDRs) and other non-pharmaceutical interventions (NPIs) such as face masks or hand hygiene precautions, computed by assuming $R_0$=3.0, as estimated for historical lineages of SARS-CoV-2 in Italy [9,10,11].
- $\theta_{\text{Alpha}}$ is a coefficient representing the transmissibility increase of the Alpha variant with respect to historical lineages. We assumed $\theta_{\text{Alpha}} = 0.5$ [12-16].
- $(1-\varphi)$ is a coefficient representing the reduction in the per-contact transmission probability ascribable to preventive transmission measures such as face masks or hand hygiene precautions. We assumed $\varphi = 20\%$ [9,10].
- $\delta(t) \in \{0,1\}$ is a scaling factor representing the proportion of pre-pandemic contacts that are active at time $t$.
- $r_a$ is the relative susceptibility to SARS-CoV-2 infection at age $a$. We sample susceptibility profiles from the posterior distribution estimated in [4], having mean values $r_a$=0.58 (95%CI 0.34-0.98) under 15 years of age; $r_a$ =1 between 15 and 64 years; and $r_a$=1.65 (95%CI 1.03-2.65) above 64 years.
- $C_{a,\tilde{a}}$ represents the age-group-specific contact matrix, as estimated before the SARS-CoV-2 pandemic [3], whose entries describe the mean numbers of persons in age group $\tilde{a}$ encountered by an individual of age group a in an average day.

- $\pi$ represents the relative infectiousness of SARS-CoV-2 cases among vaccinated compared to unvaccinated. We assumed $\pi = 0.5$ [5,6].
- $N_{\tilde{a},c}$ represents the number of individuals in the population class {ã,c}.

For all infectious compartments, the average duration of infectiousness $(1/\gamma)$ is set equal to the average generation time (6.6 days) [9]. The model described by the system of ordinary differential equations above is implemented through a stochastic discrete-time model with a time step $\tau = \frac{1}{4} day$.

At each time *t*, the number of unvaccinated individuals in the population class {a,c} (red compartments in Figure S1) who will receive a first vaccine dose is determined as a fraction $z_{a,c}(t)$ of the corresponding population:

$$z_{a,c}(t) = \frac{d_{a,c}(t)}{S_{a,c}(t) + I_{a,c}(t) + R_{a,c}(t)}$$

where $d_{a,c}(t)$ represents the number of first vaccine doses administered to individuals of the population class {a,c} at time t. The value of $d_{a,c}(t)$ is inferred from data on the daily number of first doses administered by age group in Italy between the start of vaccination (December 27, 2020) and the end of the simulated period (June 30, 2021) [17]. Details are reported in Section 1.4.

The vaccination rate $\alpha_{a,c}(t)$ associated to the probability $z_{a,c}(t)$ in the differential equations model can be computed trough the following equation

$$z_{a,c}(t) = 1 - e^{-\alpha_{a,c}(t)\tau}.$$

We assume that the first dose becomes effective on average after $1/\omega_1 = 14$ days from its administration [18], that the second dose is administered on average 42 days after the first dose (i.e. $1/\omega_1 + 1/\omega_2 = 42$ days and $1/\omega_2 = 28$ days) [19,20], and that the 2nd dose becomes effective after $1/\omega_3 = 7$ days from its administration [18]. In the baseline analysis, we assume that the protection gained after both infection and completion of the vaccination schedule lasts on average two years, i.e. $1/\upsilon_R = 1/\upsilon_V = 730$ days [7,8].

Vaccinated individuals $V_{i,a,c}$ at any stage of protection *i* can develop breakthrough infection with a susceptibility reduced by a factor $1 - VE_{i,a}^{inf}$, where $VE_{i,a}^{inf}$ represents the age-specific vaccine efficacy associated to the *i-th* stage of protection. We assume that the vaccine efficacy is the same in individuals with and without comorbidities. Vaccinated individuals for whom vaccine protection has waned ($V_{5,a,c}$) are fully susceptible to infection as unvaccinated individuals (i.e. $VE_{5,a}^{inf} = 0$ for all ages *a*). Details on the age-specific vaccine efficacy against infection are reported in Section 1.5.

#### 1.2 Reproducing the SARS-CoV-2 epidemic trajectory in Italy

The reproduction number associated to the dynamical system considered can be computed as the dominant eigenvalue of the Next Generation Matrix (NGM) [21,22,23], defined as:

$$NGM = \frac{\beta(1+\theta_{\text{Alpha}})(1-\varphi)\,\delta(t)}{\gamma} \begin{pmatrix} B_{a,\tilde{a}}^{V_0,IV_0} & B_{a,\tilde{a}}^{V_0,IV_1} & B_{a,\tilde{a}}^{V_0,IV_2} & B_{a,\tilde{a}}^{V_0,IV_3} & B_{a,\tilde{a}}^{V_0,IV_4} & B_{a,\tilde{a}}^{V_0,IV_5} \\ B_{a,\tilde{a}}^{V_1,IV_0} & B_{a,\tilde{a}}^{V_1,IV_1} & B_{a,\tilde{a}}^{V_1,IV_2} & B_{a,\tilde{a}}^{V_1,IV_3} & B_{a,\tilde{a}}^{V_1,IV_4} & B_{a,\tilde{a}}^{V_1,IV_5} \\ B_{a,\tilde{a}}^{V_2,IV_0} & B_{a,\tilde{a}}^{V_2,IV_1} & B_{a,\tilde{a}}^{V_2,IV_2} & B_{a,\tilde{a}}^{V,IV_3} & B_{a,\tilde{a}}^{V_2,IV_4} & B_{a,\tilde{a}}^{V_2,IV_5} \\ B_{a,\tilde{a}}^{V_3,IV_0} & B_{a,\tilde{a}}^{V_3,IV_1} & B_{a,\tilde{a}}^{V_3,IV_2} & B_{a,\tilde{a}}^{V_3,IV_3} & B_{a,\tilde{a}}^{V_3,IV_4} & B_{a,\tilde{a}}^{V_3,IV_5} \\ B_{a,\tilde{a}}^{V_4,IV_0} & B_{a,\tilde{a}}^{V_4,IV_1} & B_{a,\tilde{a}}^{V_4,IV_2} & B_{a,\tilde{a}}^{V_4,IV_3} & B_{a,\tilde{a}}^{V_4,IV_4} & B_{a,\tilde{a}}^{V_4,IV_5} \\ B_{a,\tilde{a}}^{V_5,IV_0} & B_{a,\tilde{a}}^{V_5,IV_1} & B_{a,\tilde{a}}^{V_5,IV_2} & B_{a,\tilde{a}}^{V_5,IV_3} & B_{a,\tilde{a}}^{V_5,IV_4} & B_{a,\tilde{a}}^{V_5,IV_5} \end{pmatrix} \quad (1)$$

Each block $B_{a,\tilde{a}}^{V_i,IV_j}$ describes the contribution to the transmission of age-specific interactions between susceptible individuals in compartment $V_i$ and infectious individuals in the compartment $IV_j$, where $IV_j = I$ for j=0, while $IV_j$ represents individuals infected in the j-stage of vaccine protection for j>0. For simplicity, we denote here the class of unvaccinated individuals with $V_0$, while $V_1, V_2, V_3, V_4$ and $V_5$ denote vaccinated individuals in the different stages of vaccine protection.
Specifically, the explicit computation of the NGM starting from model equations yields:

$$B_{a,\tilde{a}}^{V_i,IV_j} = r_a C_{a,\tilde{a}} \left[1 - VE_{i,a}^{inf}\right] \chi_{IV_j} \frac{N_{\tilde{a}}^{V_j}(t)}{N_{\tilde{a}}}$$

where:

- $VE_{i,a}^{inf}$ is the vaccine efficacy against infection for individuals of age $a$ with vaccination status $V_i$ (with $VE_{0,a}^{inf}$ = 0 by definition).
- $\chi_{IV_j}$ is the relative infectiousness of SARS-CoV-2 cases among vaccinated compared to unvaccinated (equal to $\pi$ for vaccinated compartments, and 1 for the unvaccinated).
- $N_{\tilde{a}}^{V_j}(t)$ is the number of individuals of age $\tilde{a}$ with vaccination status $V_j$ at time $t$
- $N_{\tilde{a}}$ represents the total population of age $\tilde{a}$.

The distribution of the scaling factor shaping SARS-CoV-2 transmissibility for historical lineages ($\beta$) under full resumption of pre-pandemic contacts ($\delta(t) = 1$) and in the absence of preventive measures ($\varphi = 0$ ), can be computed analytically from Equation 1 given the distribution of the age-specific susceptibility profile ($r_a$), the distribution of the bootstrapped contact matrix ($C_{a,\tilde{a}}$), the value of $\gamma$, the value of $\theta_{Alpha}$, and assuming $R_0$ = 3.0, as estimated in Italy in the early phase of the pandemic [9,10].

To reproduce the epidemic trajectory observed in Italy between January and June 2021 we use Equation 1 to recalibrate at each time step ($\tau = \frac{1}{4} day$) the value of $\delta(t)$. In particular, the selected value of $\delta(t)$ will be the one that will make the model's reproduction number (recomputed after updating the $N_{\tilde{a}}^{V_j}(t)$ to current state variables of the model) match the corresponding value of the net reproduction number as estimated on the same day from epidemic curves collected by the national integrated surveillance system [10,24] (reported in Figure S2).

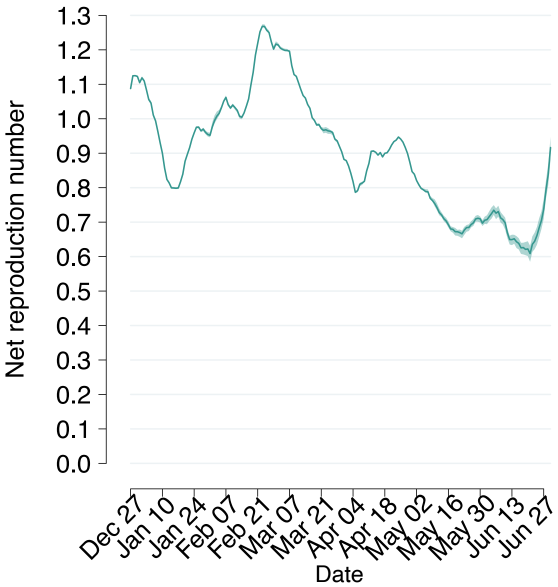

**Figure S2.** Estimates of the net reproduction number Rt the net reproduction number as obtained from epidemic curves collected by the National Integrated Surveillance System [24]

Results discussed in the main text and in the following sections were obtained by running 300 simulations, sampling at each run a different value from the joint distribution of the transmission coefficient $\beta$, the bootstrapped contact matrices $C_{a,\tilde{a}}$ and the relative susceptibility by age $r_a$.

#### 1.3 Model initialization

The model is used to simulate SARS-CoV-2 infection and vaccination dynamics in Italy between the start of the vaccination campaign (December 27, 2020) and June 30, 2021.

The population by age was initialized according to the Italian age structure in 2020 [25] and statistics on underlying conditions, among which chronic respiratory disease, cardio-cerebrovascular disease, hypertension and diabetes [26] (Figure S3).

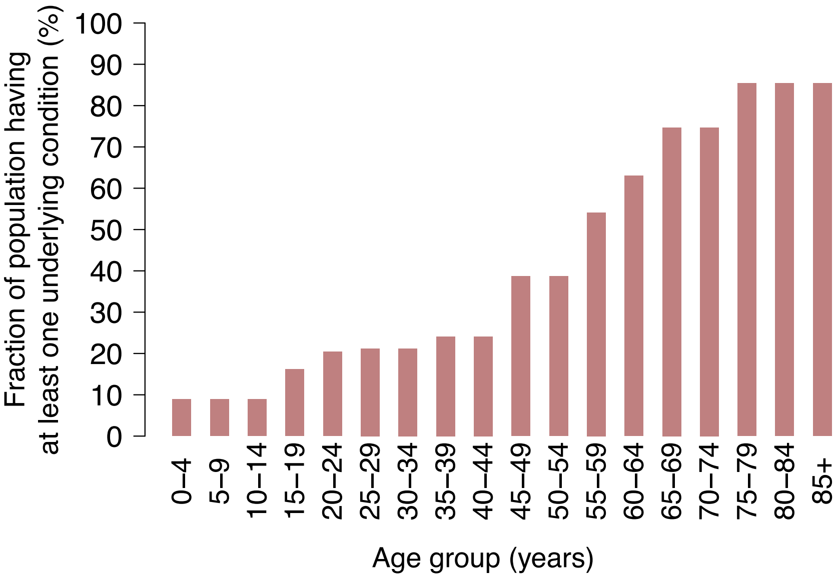

**Figure S3. Fraction of the population having at least one underlying condition by age (%) [26]**

<u>Fraction of initially immune individuals</u>. The spread of SARS-CoV-2 in Italy throughout 2020 has been characterized by marked regional heterogeneities and estimates for the fraction of immune population in Italy by the end of December 2020 are not available. We considered three different Italian regions, that are representative of different levels of SARS-CoV-2 circulation:

- Lombardy, the first and hardest hit region in 2020;
- Lazio, characterized by an intermediate circulation of SARS-CoV-2 in 2020;
- Campania, characterized by low circulation of SARS-CoV-2 in 2020.

For each region, we adapted a previously published model [1] to obtain estimates of the age-specific fraction of individuals infected with SARS-CoV-2 by December 27, 2020. Estimates obtained for Lazio were used to initialize the fraction of recovered population at the national level in the baseline analysis ("intermediate" immunity scenario), and those obtained for the two other regions were used in sensitivity analyses ("low" immunity: Campania; "high" immunity: Lombardy). For the baseline analysis (intermediate scenario), we obtained that about 16% of the overall population was infected with SARS-CoV-2 prior to the introduction of vaccination (Figure S4), while for the low and high immunity scenarios these fractions are 9% and 23%, respectively (Figure S5).

Under the assumption of natural immunity lasting on average $1/\upsilon_R$, we estimated the proportion $f$ of recovered individuals that would have lost natural immunity by December 27, 2020. To this aim, we consider the time series of daily SARS-CoV-2 cases notified in Italy throughout 2020 [27] and, for each individual case $i$ notified on day $t$, we determined if his/her natural immunity has waned by sampling the duration from an exponential distribution with average $1/\upsilon_R$.

Assuming a baseline average value for the duration of natural immunity of 2 years, we obtained that a fraction $f$=8.9% of individuals recovered from SARS-CoV-2 by the end of 2020 may have lost natural immunity by that time (light blue in Figure S4-S5).

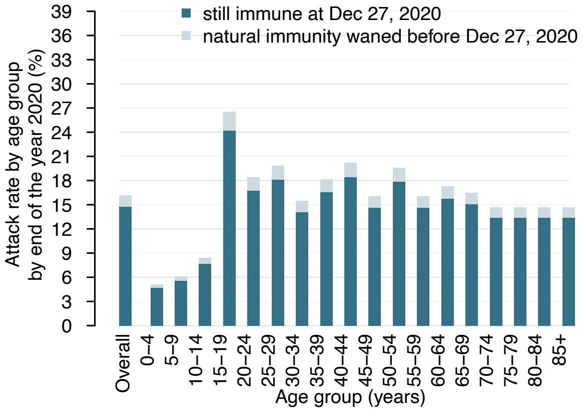

**Figure S4. Intermediate immunity scenario.** Overall and age-specific fraction of population with natural immunity to SARS-CoV-2 in the intermediate immunity scenario at the beginning of the vaccination campaign [1].

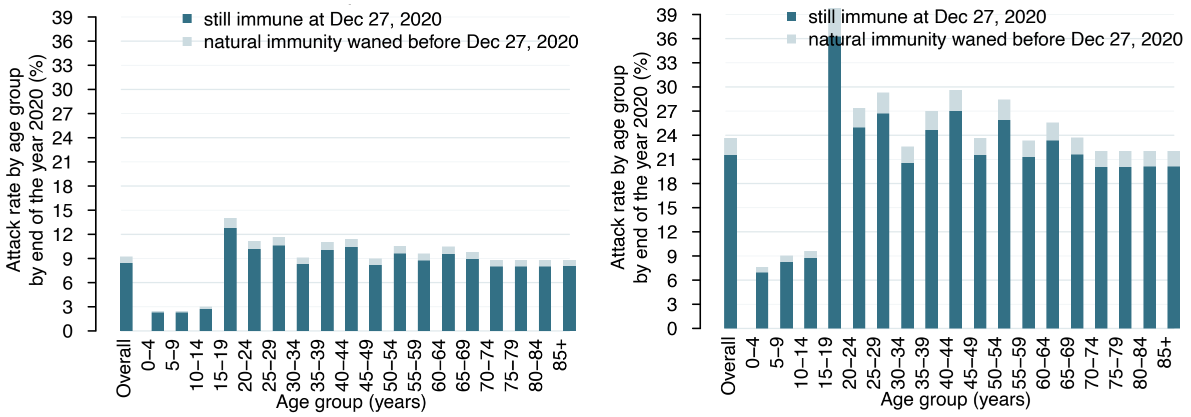

**Figure S5. Low and high immunity scenarios.** Left: Overall and age-specific fraction of population with natural immunity to SARS-CoV-2 in the low immunity scenario at the beginning of the vaccination campaign [1]. Right: The same as left, but for the high immunity scenario.

**Initially infectious individuals.** The number of infected individuals at the beginning of simulations (December 27, 2020) was determined in such a way to match the average daily number of notified cases reported in the week of simulations start (around 15,000) [27]. The reporting rate $\rho$ for SARS-CoV-2 infections at the end of 2020 was estimated as

$$\rho = \varepsilon / CFR$$

where $\varepsilon$ is the infection fatality ratio estimated for Italy [28] and CFR is the case fatality ratio among SARS-CoV-2 cases reported in the last three weeks of December, assuming a delay between diagnosis and death of about 3 weeks [29]; thus, CFR=$M/C$ where C is the cumulative number of cases reported in the last 3 weeks of December 2020 and M represents the number of deaths reported in the first 3 weeks of January 2021 [30]. The resulting value for the reporting rate is 41.2%.

#### 1.4 Computation of age-specific vaccination rates over time

The rollout of the two-dose vaccination campaign is modeled using detailed data on the daily age-specific number of first doses administered over the considered period [17]. In the model, second doses are assumed to be administered to all individuals who have been vaccinated with one dose after an average of 42 days since the first dose. As shown in Figure S6, the model well reproduces the observed scale-up of the daily vaccination capacity occurred in Italy in the first half of the year 2021. The priority order of the Italian vaccination campaign was based on the WHO SAGE roadmap [31, 32] prioritizing high-risk population age segments, i.e. over 80 years of age and essential workers (e.g. health care workers and teachers) and then progressively targeting younger age groups. In the absence of data regarding the presence or absence of underlying conditions in individuals targeted by vaccination, we assumed to distribute the daily doses proportionally to the prevalence of underlying conditions in the age group considered. The evolution of vaccination coverage, by age and overall, as observed in the first half of the year 2021 is shown in Figure S6.

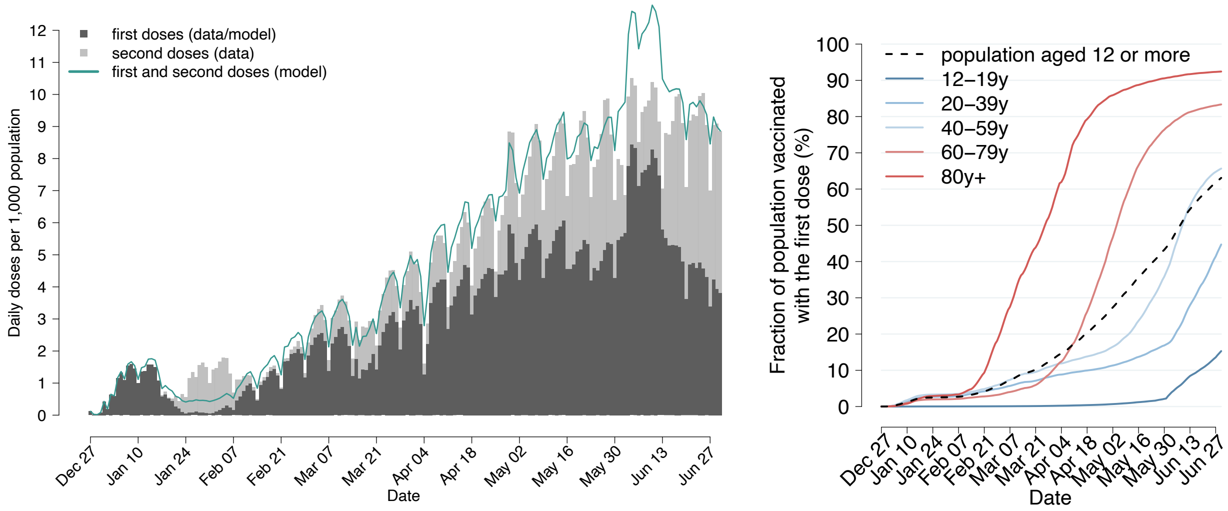

**Figure S6.** Left: daily vaccination capacity in Italy (number of doses per 1,000 population) as observed [17] (grey bars) and as estimated by the model (solid line) between December 27, 2020, and June 30, 2021. Right: first dose vaccination coverage overall (black dotted line) and by age group (solid lines) as observed in Italy between December 27, 2020, and January 30, 2021 [17].

#### 1.5 Vaccine efficacy against infection and death

In the first half of 2021 about 51 million vaccine doses have been administered in Italy, about 71% of which were Pfizer- BioNTech, 17% AstraZeneca, 10% Moderna and 2% Janssen [17].

Current evidence suggests an efficacy against infection with the Alpha variant of about 62% after two doses of AstraZeneca [33]. Estimates available for Pfizer-BioNTech suggest an efficacy against infection with Alpha of about 89% after two doses [34]. We inferred the corresponding vaccine efficacies after one dose of vaccine by assuming a 11% relative reduction compared to the efficacy observed after two doses [35, 36] (Table S1).

**Table S1**. Vaccine efficacy against symptomatic and asymptomatic infection

| | 1 DOSE | 2 DOSES |
|---|---|---|
| **Pfizer-BioNTech (mRNA)** | 79%* | 89% [34] |
| **AstraZeneca (viral vector)** | 55%* | 62% [33] |

*inferred

Using values reported in Table S1, we estimated an age-specific vaccine efficacy against infection by weighting the efficacy of a specific vaccine type (mRNA vs. viral vectors) by the number of vaccines of that type administered to each age group in the first half of 2021 [17] (Figure S7). We assume for the Moderna vaccine (mRNA type) the same vaccine efficacy of Pfizer-BioNTech and for Janssen (viral vector type) the vaccine same vaccine efficacy of AstraZeneca (Table S1). Obtained age-specific estimates are in good agreement with estimates obtained for Italy [37] and range between 70.6% and 78.8% after the first dose (average 75.5%) and between 79.4% and 88.7% after 2 doses (average: 84.9%).

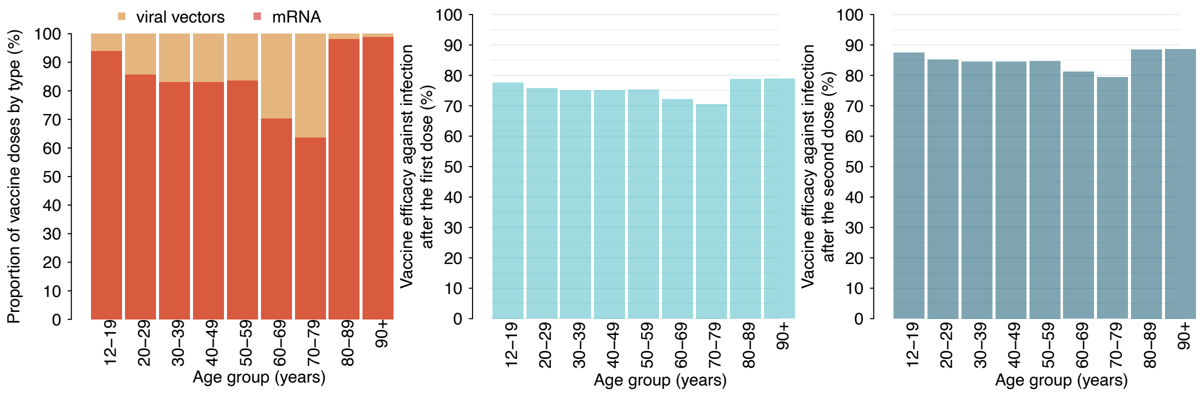

**Figure S7.** Left: Age-specific proportion of vaccine doses by vaccine type administered in the first half of 2021. mRNA vaccines administered in Italy include Pfizer-BioNTech and Moderna, while viral vector vaccines include AstraZeneca and Janssen. Center: Estimated age-specific vaccine efficacy against infection after the first dose (%). Right: Estimated age-specific vaccine efficacy against infection after the second dose (%).

Available estimates suggest a marked reduction in the risk of death in breakthrough infections: about 80.3% in vaccinated with one dose and about 96.4% in vaccinated with two doses [37]. These values include the reduced risk of SARS-CoV-2 infection in vaccinated compared to unvaccinated. To compute the infection fatality rate in breakthrough infections at different stages of vaccine protection $V_j$, we estimate an age-specific scaling coefficient $\mu_a^{V_j}$ representing the risk of death given SARS-CoV-2 breakthrough infection in each stage $V_j$ relative to infection in unvaccinated individuals:

$$\mu_a^{V_j} = \frac{1 - VE_{j,a}^{\text{inf}}}{1 - VE_j^{\text{death}}}$$

Where $VE_j^{\text{death}}$is set to 0 for unprotected individuals (j=0, 1, 5), to 80.3% for partially protected individuals (j=2, 3), and 96.4 for fully vaccinated individuals (j=4).

#### 1.6 Model outputs

The main model outcomes are the age-specific number of new infections per day in the subpopulation $\mathrm{i}_{\mathrm{a,c}}^{V_j}(\mathrm{t})$ for each vaccination stage *j* (including the unvaccinated, j=0); and the scaling factor $\delta(\mathrm{t})$ tuning the proportion of pre-pandemic contacts necessary reproduce the daily official estimates of Rt.

Additional model outcomes are:

- the number of SARS-CoV-2 confirmed cases between December 27, 2020, and June 30, 2021 and cumulative number of COVID-19 deaths over the same period;
- the immunity profile of the Italian population on June 30, 2021;
- estimates of the effective reproduction number (i.e., under the assumption of $\delta(\mathrm{t})$=1).

For each model outcome, we report the mean values and 95% confidence intervals across stochastic simulations.

**SARS-CoV-2 confirmed cases and COVID-19 deaths.**

The daily number of SARS-CoV-2 confirmed cases is obtained from the daily infections by assuming a reporting ratio ($\rho$) of 41.2% (see Section 1.3). The daily number of deaths in each subpopulation {a,c} with vaccination status $V_j$ is obtained by applying the age-specific infection fatality ratio estimated for each population class ($\varepsilon_{\mathrm{a,c}}$) to daily infections, considering a delay of 3 weeks ($\tau_{\mathrm{D}}$) between symptom onset and death [29], and rescaled by a factor $(1 - q)$ to account for improvement in COVID-19 treatment. Specifically,

$$\mathrm{D}_{\mathrm{a,c}}^{V_j}(\mathrm{t}) = \mu_{\mathrm{a}}^{V_j}\,(1 - q)\varepsilon_{\mathrm{a,c}}\,\mathrm{i}_{\mathrm{a,c}}^{V_j}(\mathrm{t} - \tau_{\mathrm{D}})$$

where $\mu_a^{V_j}$ is the relative risk of death in breakthrough infections, as computed in Section 1.5. To match the cumulative number of deaths detected by the surveillance system over the considered period, we obtain a reduction of mortality $q$ by 20%.

The age-specific IFR for each population class ($\varepsilon_{\mathrm{a,c}}$) was estimated as:

$$\varepsilon_{a,c} = \psi_c \cdot \varepsilon_a = \nu \cdot \varepsilon_c \cdot \varepsilon_a$$

where

- $\varepsilon_c$ denotes the overall IFR in class c, as estimated from Lombardy data [28].
- $\varepsilon_a$ denotes the age-specific IFR as estimated independently of the presence of underlying conditions [28].
- the scale factor $\nu$ is determined in such a way to minimize the root mean square error between $\varepsilon_a$ and $\tilde{\varepsilon}_a = \sum_c P_{a,c} \cdot \varepsilon_{a,c}$, and $P_{a,c}$ denotes the proportions of individuals of age a in class c in the Italian demographics [26].

Eventually, we obtain $\psi_{without}$=0.20 for individuals without underlying conditions and $\psi_{with}$=1.16 for individuals with underlying conditions, consistent with the overall relative risk $\frac{\psi_{with}}{\psi_{without}}$ = 5.8 estimated in [28].

**Effective reproduction number.** For each time t, the effective reproduction number is computed as the dominant eigenvalue of the Next Generation Matrix defined in Equation (1) by setting $\delta(t) = 1$, to account for complete resumption of pre-pandemic contacts (i.e., in absence of interventions and behavioral change).

#### 1.7 No vaccination scenario

To evaluate the impact of the vaccination campaign, we need to define a suitable counterfactual scenario. Simply removing vaccination from the model and letting the epidemics evolve in absence of interventions would be an unrealistic scenario in terms of public health. Therefore, we chose as counterfactual a scenario where governmental restrictions and individual behavior would result in the same epidemic trajectory as the one observed in the presence of vaccination. We set the daily number of first doses $d_{a,c}(t)$ to 0 for all t, and recalibrated at each time step the value of $\delta(t)$ necessary to match the time-series of the net reproduction number estimated from surveillance data (Figure S2) [10, 24]. Then, we compared the obtained estimates of social contacts, deaths, and effective reproduction number with those of the main analysis.

#### 1.8 Future vaccination scenarios for the Delta variant

**Summer 2021**. We assess the combined effect of the replacement of Alpha by the Delta variant and of the progression of the vaccination campaign in July and August.
To this aim, we estimate the reproduction number on September 7, 2021, accounting for the progression of vaccination and assuming that the proportion of active pre-pandemic contacts has not changed during summer 2021.
The reproduction number can be computed through Equation 1 by:

- updating the SARS-CoV-2 immunity profile $\frac{N_{\tilde{a}}^{V_j}(t)}{N_{\tilde{a}}}$ to account for the increment in age-specific vaccination coverage between June 30, 2021, and September 7, 2021;
- setting the proportion of active pre-pandemic contacts to the value estimated on June 30, 2021, i.e. $\delta(t) = \delta(t^*)$ where $t^*$ corresponds to June 30, 2021.
- adding a multiplying scale factor $(1 + \theta_{Delta})$ when assuming that the Delta variant is dominant. The baseline value assumed for $\theta_{Delta}$ is 50% [38-40].

This estimate neglects the increase in natural immunity due to SARS-CoV-2 infection occurring during the summer. This choice is supported by the low viral circulation in Italy in the summer, with 322,191 reported cases between July 1 and September 7, 2021, corresponding to about 1.3% of the total Italian population being infected after accounting for underreporting. By comparison, about 13.3% of all Italians have been fully vaccinated over the same period. We also did not mechanistically model the transmission dynamics of SARS-CoV-2 during the summer months because of the complexities arising from the co-circulation of two distinct strains. For the purpose of our estimates, the specific dynamics of COVID-19 transmission in the summer is irrelevant. In fact, the reproduction numbers estimated here only depend on the profile of population immunity at the considered date.

**Future vaccination scenarios.** We provide estimates on the reproduction number to be expected for the Delta variant under a set of vaccination scenarios $\Omega$, corresponding to different improvements of the vaccination coverage with respect to September 7, 2021. Vaccination scenarios $\Omega$ are defined as follows: let $\mathrm{Cov}^*(a)$ be the vaccination coverage achieved by September 7, 2021 in age group $a$, we define the age-specific vaccination coverage in age group $a$ under scenario $\Omega$ as:

$$\mathrm{Cov}(a,\Omega)=\begin{cases}0 & \text{if } a<a_{MIN}\\ \max\{\mathrm{Cov}^{*}(a)\,,\Omega\} & \text{if } a\geq a_{MIN}\end{cases}$$

where $a_{MIN}$ is the minimum age to which the vaccine is administered (12 years in the baseline analysis, 5 years when considering a pediatric vaccine). We explored values of Ω between 60% and 100% with incremental steps of 5%.

For each vaccination scenario $\Omega$, we compute the reproduction number associated to different social contact levels $\delta^{*}$ (between 0% and 100% of pre-pandemic contacts, with incremental steps of 5%) as the dominant eigenvalue of the Next Generation Matrix defined as:

$$NGM(\delta^{*},\Omega)=\frac{\beta\left(1+\theta_{\mathrm{Alpha}}\right)(1+\theta_{\mathrm{Delta}})(1-\varphi)\,\delta^{*}}{\gamma}\begin{pmatrix} B_{a,\tilde{a}}^{V_0,IV_0}(\Omega) & B_{a,\tilde{a}}^{V_0,IV_1}(\Omega) & B_{a,\tilde{a}}^{V_0,IV_2}(\Omega) & B_{a,\tilde{a}}^{V_0,IV_3}(\Omega) & B_{a,\tilde{a}}^{V_0,IV_4}(\Omega) & B_{a,\tilde{a}}^{V_0,IV_5}(\Omega)\\ B_{a,\tilde{a}}^{V_1,IV_0}(\Omega) & B_{a,\tilde{a}}^{V_1,IV_1}(\Omega) & B_{a,\tilde{a}}^{V_1,IV_2}(\Omega) & B_{a,\tilde{a}}^{V_1,IV_3}(\Omega) & B_{a,\tilde{a}}^{V_1,IV_4}(\Omega) & B_{a,\tilde{a}}^{V_1,IV_5}(\Omega)\\ B_{a,\tilde{a}}^{V_2,IV_0}(\Omega) & B_{a,\tilde{a}}^{V_2,IV_1}(\Omega) & B_{a,\tilde{a}}^{V_2,IV_2}(\Omega) & B_{a,\tilde{a}}^{V,IV_3}(\Omega) & B_{a,\tilde{a}}^{V_2,IV_4}(\Omega) & B_{a,\tilde{a}}^{V_2,IV_5}(\Omega)\\ B_{a,\tilde{a}}^{V_3,IV_0}(\Omega) & B_{a,\tilde{a}}^{V_3,IV_1}(\Omega) & B_{a,\tilde{a}}^{V_3,IV_2}(\Omega) & B_{a,\tilde{a}}^{V_3,IV_3}(\Omega) & B_{a,\tilde{a}}^{V_3,IV_4}(\Omega) & B_{a,\tilde{a}}^{V_3,IV_5}(\Omega)\\ B_{a,\tilde{a}}^{V_4,IV_0}(\Omega) & B_{a,\tilde{a}}^{V_4,IV_1}(\Omega) & B_{a,\tilde{a}}^{V_4,IV_2}(\Omega) & B_{a,\tilde{a}}^{V_4,IV_3}(\Omega) & B_{a,\tilde{a}}^{V_4,IV_4}(\Omega) & B_{a,\tilde{a}}^{V_4,IV_5}(\Omega)\\ B_{a,\tilde{a}}^{V_5,IV_0}(\Omega) & B_{a,\tilde{a}}^{V_5,IV_1}(\Omega) & B_{a,\tilde{a}}^{V_5,IV_2}(\Omega) & B_{a,\tilde{a}}^{V_5,IV_3}(\Omega) & B_{a,\tilde{a}}^{V_5,IV_4}(\Omega) & B_{a,\tilde{a}}^{V_5,IV_5}(\Omega)\end{pmatrix}$$

where

$$B_{a,\tilde{a}}^{V_i,IV_j}(\Omega)=r_a C_{a,\tilde{a}}\left[1-VE_{i,a}^{inf}\right]\chi_{IV_j}\frac{N_{\tilde{a}}^{V_j}(\Omega)}{N_{\tilde{a}}}$$

and $N_{\tilde{a}}^{V_j}(\Omega)$ represents the number of individuals of age $\tilde{a}$ with vaccination status $V_j$ according to vaccination scenario $\Omega$.

As a baseline, we assume a transmissibility increase for the Delta variant with respect to Alpha ($\theta_{\mathrm{Delta}}$) equal to 50% [38-40]. Alternative values of $\theta_{\mathrm{Delta}}$ (i.e. 25% and 75%) are explored as sensitivity analyses.

### 2. Sensitivity analyses

#### 2.1 Description

We assess the robustness of our results with respect to alternative assumptions on:

- the average duration of natural immunity ($1/\upsilon_R$);
- the average duration of vaccine-induced immunity ($1/\upsilon_V$);
- the relative infectiousness of SARS-CoV-2 breakthrough infections ($\pi$);
- the natural immunity level at model initialization (end of December 2020)

Table S2 summarizes the main model parameters and assumptions in the baseline and in alternative sensitivity analyses.

**Table S2.** Description of key parameters and assumption used in the model and in alternative scenarios

| Parameter description | Baseline | Alternative | Source |
|---|---|---|---|
| **Epidemiological** | | | |
| Generation time ($1/\gamma$) | 6.6 days | - | [9] |
| Age-specific susceptibility to infection ($r_a$) | $r_a = 0.58$ (95%CI 0.34-0.98) for $a < 15$ years; $r_a = 1$ for $15 \leq a < 65$ years; $r_a = 1.65$ (95%CI 1.03-2.65) when $a \geq 65$ years | - | [4] |
| Duration of immunity after SARS-CoV-2 infection ($1/\upsilon_R$) | 2 years | • 1 year<br>• 10 years | [7] |
| Age-group specific contact matrix ($C_{a,\tilde{a}}$) | Contact matrix estimated for Italy before the COVID-19 pandemic | - | [3] |
| Initially immune individuals by age | intermediate immunity scenario (average ~15%) | • low immunity scenario (average ~8%)<br>• high immunity scenario (average ~21%) | Estimated [1] (see Section 1.3) |
| SARS-CoV-2 reporting ratio ($\rho$) | 41.2% | - | Estimated (see Section 1.3) |
| **Vaccination** | | | |
| Number of doses | 2 | - | |
| Delay between 1st dose and achievement of vaccine efficacy | 14 days | - | [18] |
| Interval between 1st and 2nd dose | 42 days | - | [19,20] |
| Delay between 2nd dose and achievement of vaccine efficacy | 7 days | - | [18] |
| Efficacy against infection immediately after 1st dose ($VE_{1,a}^{inf}$) | 0% for all ages | - | |
| Full efficacy against infection of 1st dose ($VE_{2,a}^{inf}$) | Age-specific: average 75.5% (range 70.6%-78.8%) | - | Estimated (see Section 1.5) |
| Efficacy against infection immediately after 2nd dose ($VE_{3,a}^{inf}$) | Same as $VE_{2,a}$ | - | Assumed |
| Full efficacy against infection of 2nd dose ($VE_{4,a}$) | Age-specific: average 88.7% (range 79.4%-88.7 %) - see Section 1.5 | - | Estimated (see Section 1.5) |
| Efficacy against infection after waning of vaccine protection ($VE_{5,a}$) | 0% for all ages | - | Assumed |
| Relative infectiousness of SARS-CoV-2 breakthrough infections ($\pi$) | 50% | 100% | [5,6] |
| Duration of vaccine protection ($1/\upsilon_V$) | 2 years | • 1 year<br>• 10 years | [8] |

#### 2.2 Results

Figure S8-S10 shows that model results are most sensitive to the duration of natural immunity and the initial immunity profile, but that overall the estimates are quite robust. In particular, the estimate for June 30, 2021 of the average proportion of fully susceptible individuals ranges between 34% and 38% (Figure S8), that of social contacts ranges between 44% and 53% of pre-pandemic contacts (Figure S9) and that of the effective reproduction number ranges between 1.7 and 2.1 (Figure S10).
Results obtained for future vaccination scenarios suggest that, if the vaccine did not reduce the infectiousness of breakthrough infections, the social contact levels that could be achieved without causing an epidemic would be about 45-65% (depending on the coverage scenario) compared to 55%-70% for the baseline (Figure S11).
The duration of vaccine-induced protection affects very marginally results for the prospective vaccination scenarios (Figure S12), due to the fact that the majority of vaccines have been administered too recently for waning to have a major effect. On the other hand, shorter duration of natural immunity, or a lower level of natural immunity at model initialization, would result in a slightly lower level of social contacts that could be resumed without causing an epidemic (namely, 50-70% for a duration of immunity of 1 year and 50-70% for an initial immunity profile that is lower than estimated - see Figures S13-S14, left panels).

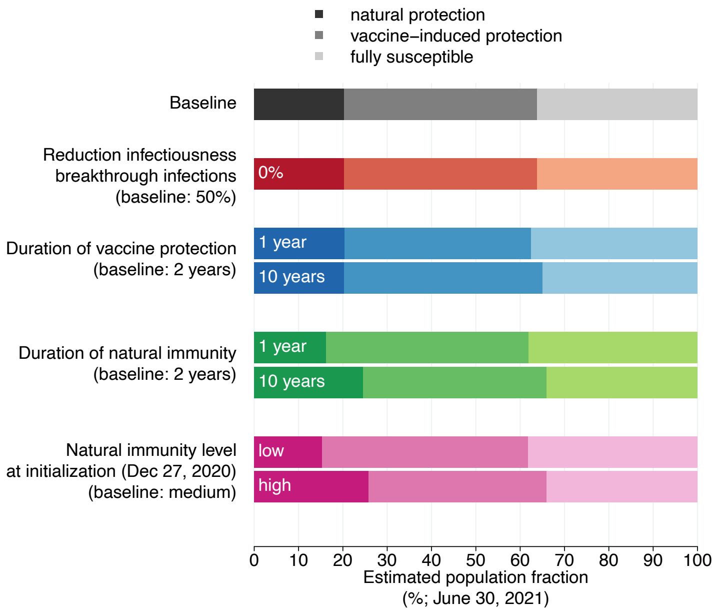

**Figure S8.** Estimated immunity profile of the overall Italian population on June 30, 2021 for the baseline analysis and for the sensitivity analyses considered. Individuals who have been infected after being vaccinated or who have been vaccinated despite still having a protection from infection are counted under the natural protection bar; individuals who have never been infected or who have lost their natural protection and were vaccinated (partially or fully) are included under the vaccine-induced protection bar; individuals who were never vaccinated nor infected, or who were infected but lost their natural protection, or who were vaccinated but lost their vaccine-induced protection are included under the fully susceptible bar.

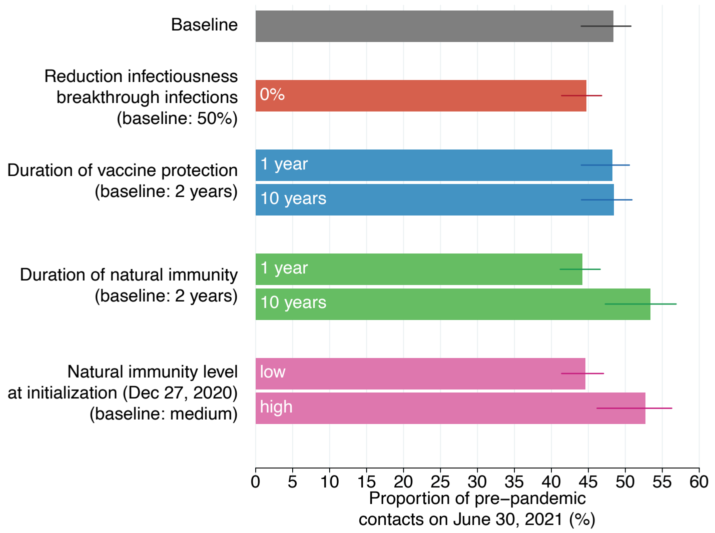

**Figure S9.** Estimated active social contacts on June 30, 2021, as a proportion of pre-pandemic contacts for the baseline analysis and for the sensitivity analyses considered. Bars: mean estimates; horizontal lines: 95% CI.

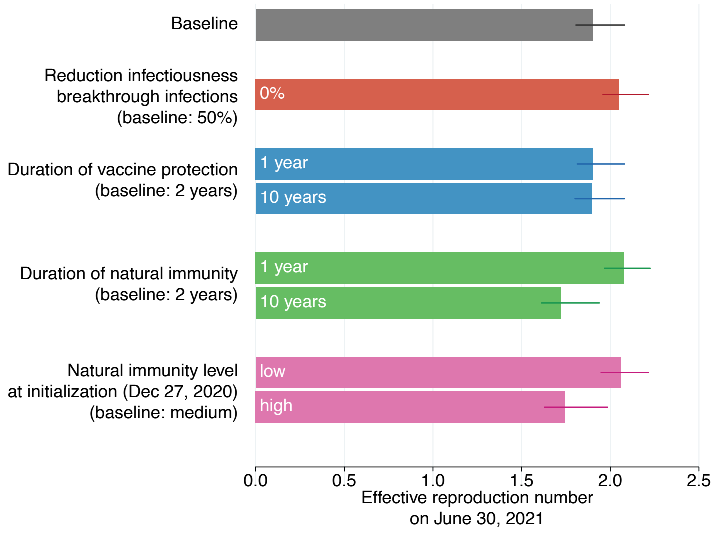

**Figure S10.** Effective reproduction number (i.e., under complete resumption of pre-pandemic contacts) on June 30, 2021 for the baseline analysis and for the sensitivity analyses considered. Bars: mean estimates; horizontal lines: 95% CI.

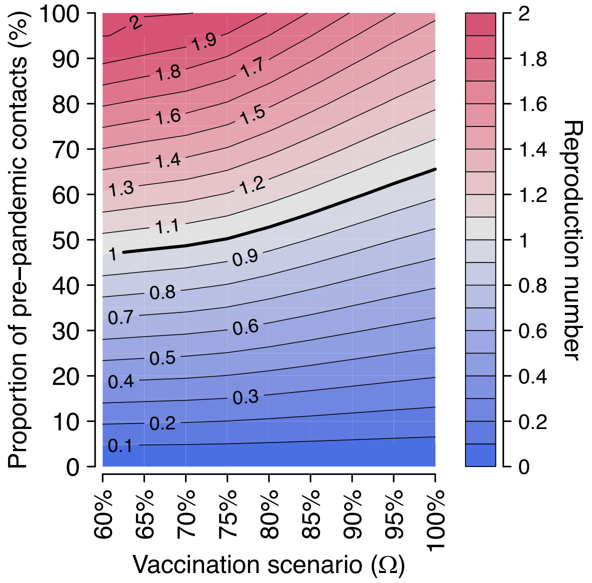

**Figure S11. Sensitivity analysis with respect to the relative infectiousness of SARS-CoV-2 breakthrough infections (π=100%).** Heatmap of the estimated reproduction number for different vaccination scenarios (x axis) and different levels of social activity (y axis). Contour lines discriminate different values of the reproduction number. The thicker contour line represents the epidemic threshold of 1.

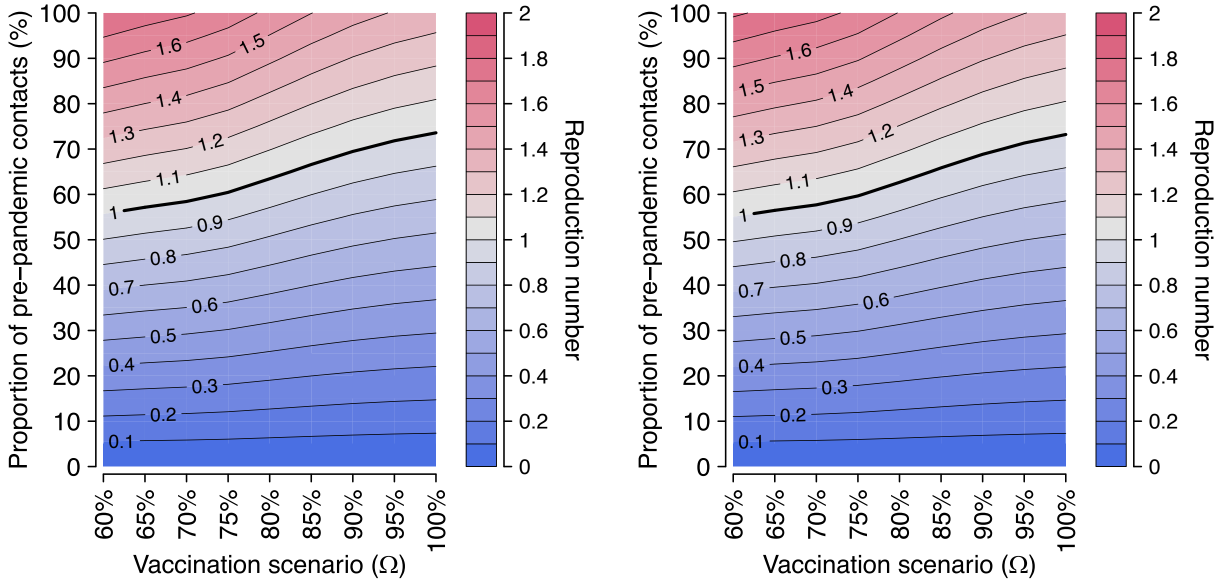

**Figure S12. Sensitivity analysis with respect to the duration of vaccine-induced immunity ($1/v_V$).** Heatmap of the estimated reproduction number for different vaccination scenarios (x axis) and different levels of social activity (y axis), under different average durations of vaccine protection: 1 year (left) and 10 years (right)

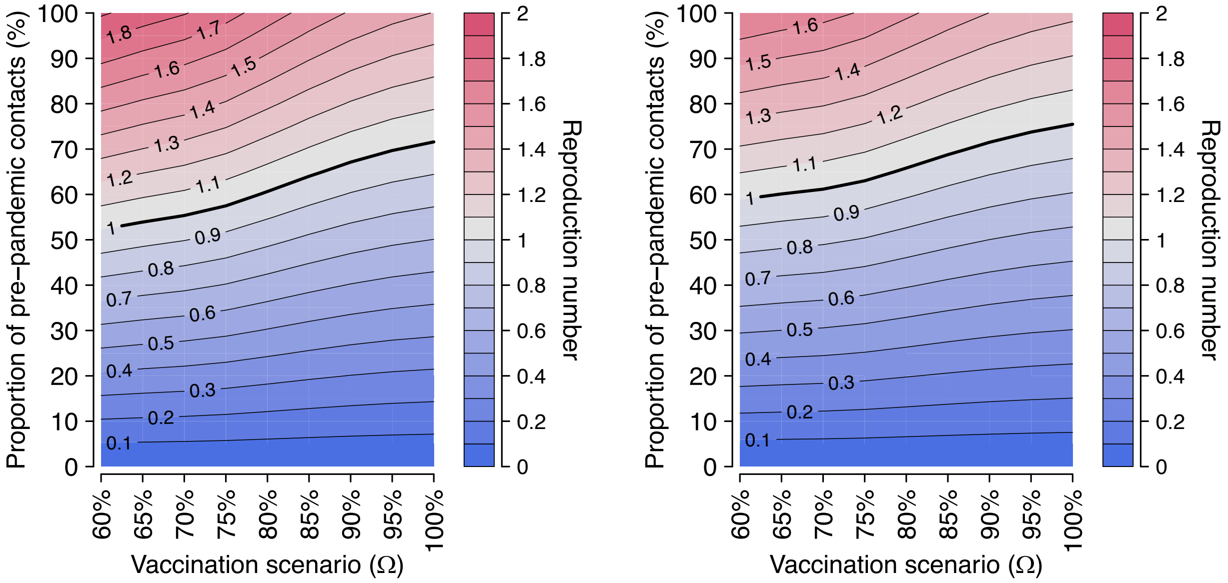

**Figure S13. Sensitivity analysis with respect to the duration of natural immunity ($1/v_R$).** Heatmap of the estimated reproduction number for different vaccination scenarios (x axis) and different levels of social activity (y axis), under different average durations of natural immunity: 1 year (left) and 10 years (right).

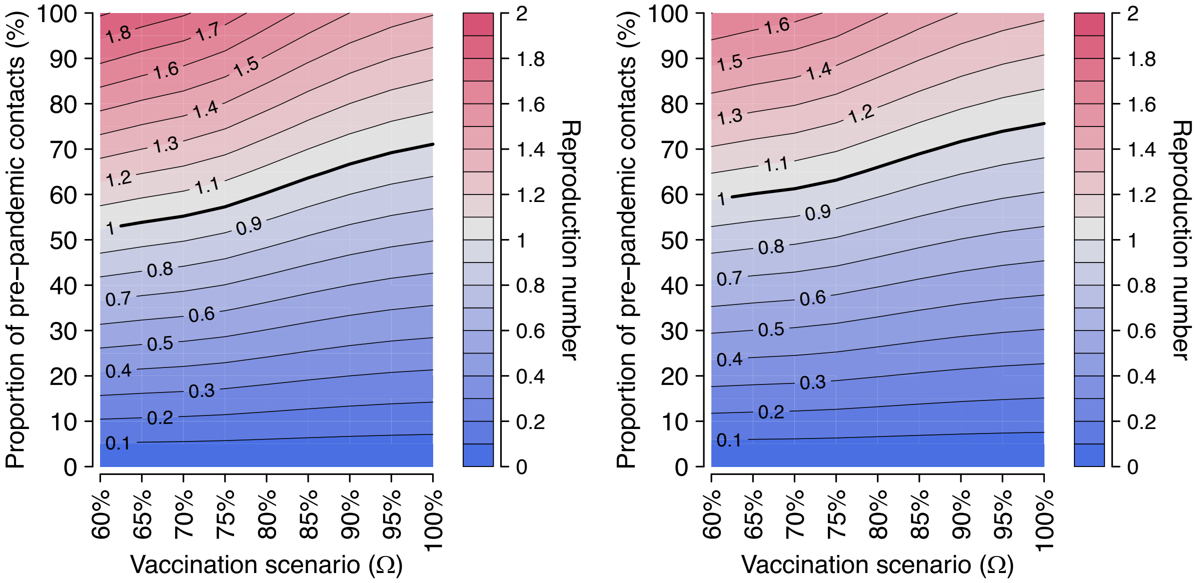

**Figure S14. Sensitivity analysis with respect to the level of initial immunity.** Heatmap of the estimated reproduction number for different vaccination scenarios (x axis) and different levels of social activity (y axis), under different initial levels of natural immunity: low immunity scenario (left) and high immunity scenario (right).
